# Assessing the performance of a serological point-of-care test in measuring detectable antibodies against SARS-CoV-2

**DOI:** 10.1101/2021.02.04.21251126

**Authors:** Peter V. Coyle, Reham Awni El Kahlout, Soha R. Dargham, Hiam Chemaitelly, Mohamed Ali Ben Hadj Kacem, Naema Hassan Abdulla Al-Mawlawi, Imtiaz Gilliani, Nourah Younes, Zaina Al Kanaani, Abdullatif Al Khal, Einas Al Kuwari, Andrew Jeremijenko, Anvar Hassan Kaleeckal, Ali Nizar Latif, Riyazuddin Mohammad Shaik, Hanan F. Abdul Rahim, Gheyath K. Nasrallah, Hadi M. Yassine, Mohamed G. Al Kuwari, Hamad Eid Al Romaihi, Patrick Tang, Roberto Bertollini, Mohamed H. Al-Thani, Laith J. Abu-Raddad

**Affiliations:** Hamad Medical Corporation, Doha, Qatar; Infectious Disease Epidemiology Group, Weill Cornell Medicine-Qatar, Cornell University, Doha, Qatar; World Health Organization Collaborating Centre for Disease Epidemiology Analytics on HIV/AIDS, Sexually Transmitted Infections, and Viral Hepatitis, Weill Cornell Medicine–Qatar, Cornell University, Qatar Foundation – Education City, Doha, Qatar; College of Health Sciences, QU Health, Qatar University, Doha, Qatar; Biomedical Research Center, Qatar University, Doha, Qatar; Department of Biomedical Science, College of Health Sciences, Member of QU Health, Qatar University, Doha, Qatar; Primary Health Care Corporation, Doha, Qatar; Ministry of Public Health, Doha, Qatar; Department of Pathology, Sidra Medicine, Doha, Qatar; Department of Population Health Sciences, Weill Cornell Medicine, Cornell University, New York, New York, USA

## Abstract

**Objective:** To investigate the performance of a rapid point-of-care antibody test, the BioMedomics COVID-19 IgM/IgG Rapid Test, in comparison with a high-quality, validated, laboratory-based platform, the Roche Elecsys Anti-SARS-CoV-2 assay.

**Methods:** Serological testing was conducted on 708 individuals. Concordance metrics were estimated. Logistic regression was used to assess associations with seropositivity.

**Results:** SARS-CoV-2 seroprevalence was 63.4% (449/708; 95% CI 59.8%-66.9%) using the BioMedomics assay and 71.9% (509/708; 95% CI 68.5%-75.1%) using the Elecsys assay. There were 62 discordant results between the two assays. One specimen was seropositive in the BioMedomics assay, but seronegative in the Elecsys assay, while 61 specimens were seropositive in the Elecsys assay, but seronegative in the BioMedomics assay. Positive, negative, and overall percent agreements between the two assays were 88.0% (95% CI 84.9%-90.6%), 99.5% (95% CI 97.2%-99.9%), and 91.2% (95% CI 88.9%-93.1%), respectively, with a Cohen’s kappa of 0.80 (95% CI 0.77-0.83), indicating excellent agreement. Excluding specimens with lower antibody titers, the agreement improved with positive, negative, and overall percent concordance of 91.2% (95% CI 88.2%-93.6%), 99.5% (95% CI 97.2%-99.9%), and 93.9% (95% CI 91.7%-95.5%), respectively, and a Cohen’s kappa of 0.87 (95% CI 0.84-0.89). Logistic regression confirmed better agreement with higher antibody titers.

**Conclusion:** The BioMedomics COVID-19 IgM/IgG Rapid Test demonstrated excellent performance in measuring detectable antibodies against SARS-CoV-2, supporting the utility of such rapid point-of-care serological testing to guide the public health responses and possible vaccine prioritization.

## Introduction

Caused by the novel severe acute respiratory syndrome coronavirus 2 (SARS-CoV-2), coronavirus disease 2019 (COVID-19) continues to present a global challenge, causing severe health, social, and economic burdens [1, 2]. Qatar experienced a large SARS-CoV-2 epidemic with a high rate of laboratory-confirmed infections at >60,000 infections per million population (6%) [3–5]. As part of the national response, public health authorities expanded serological testing for SARS-CoV-2 antibodies, for both healthcare and research purposes [6–9]. Moreover, antibody status is being deliberated as one of the criteria for COVID-19 vaccine prioritization [10].

To achieve more efficient, cost-effective, and widescale serological testing, the objective of this study was to investigate the performance of a rapid point-of-care antibody test, the BioMedomics COVID-19 IgM/IgG Rapid Test [11], in comparison with a high-quality, validated, laboratory-based assay, the Roche Elecsys Anti SARS-CoV-2 platform [12, 13], one of the most extensively used and investigated commercial platforms, having a specificity ≥99.8% [14, 15] and a sensitivity ≥89% [3, 12, 15].

## Methods

Samples included 708 residual blood serum specimens that were collected and stored between May 30 and November 18, 2020 from individuals receiving routine or other clinical care at Hamad Medical Corporation (HMC), the main provider of healthcare in Qatar, and the nationally-designated provider for COVID-19 healthcare needs.

Serological testing was performed using the Roche Elecsys Anti-SARS-CoV-2 (Roche, Switzerland) assay, a fully-automated electrochemiluminescent immunoassay [13], and the BioMedomics COVID-19 IgM/IgG Rapid Test (BioMedomics, Inc., United States of America), a lateral flow immunochromatographic assay [11].

The Roche Elecsys Anti-SARS-CoV-2 assay (hereafter “Elecsys”) uses a recombinant protein representing the nucleocapsid (N) antigen for determination of antibodies against SARS-CoV-2 [13]. Qualitative anti-SARS-CoV-2 results were generated following the manufacturer’s instructions (reactive for optical density (proxy for antibody titer [15]) cutoff index ≥1.0 vs. non-reactive for cutoff index <1.0) [13].

The BioMedomics COVID-19 IgM/IgG Rapid Test (hereafter “BioMedomics”) is a lateral flow immunoassay that contains a colloidal, gold-labeled, recombinant coronavirus antigen and a quality control antibody colloidal gold marker, two detection lines (IgG and IgM lines), and one quality control line (C) fixed on a nitrocellulose membrane [11]. Qualitative anti-SARS-CoV-2 results were generated by reading the detection line(s) [11].

Results of serological testing were subsequently linked to the national centralized SARS-CoV-2 polymerase chain reaction (PCR) testing and hospitalization database that includes records for all PCR testing and COVID-19 hospitalizations in Qatar since the start of the epidemic [16]. The database also includes the severity classification of hospitalized cases based on individual chart reviews completed by trained medical personnel using the World Health Organization (WHO) criteria [17].

Cross-tabulations of serological testing results were conducted using the Elecsys assay as the reference standard. Concordance metrics were estimated and included the positive, negative, and overall percent agreements. In addition, Cohen’s kappa statistic was estimated to measure the level of agreement, beyond chance, between the two diagnostic approaches [18]. Ranging between 0 and 1, a kappa statistic <0.40 indicates poor agreement, a value between 0.40 and 0.75 denotes fair/good agreement, and a value >0.75 signifies excellent agreement [18]. Significance was set at 5% and a 95% confidence interval (CI) was estimated for each metric.

Univariable and multivariable logistic regression were conducted to assess associations between seropositivity using the BioMedomics assay and each of the following covariates: PCR Ct value, optical density value of the Elecsys assay result, and severity of infection. Covariates with p-values ≤0.2 in the univariable regression analysis were included in the multivariable model. Covariates with p-values ≤0.05 in the multivariable analysis were regarded as strong evidence for an association with the outcome. Analyses were performed using Microsoft Excel and IBM-SPSS version 26.0.

Research methods were approved by the ethics review boards at Hamad Medical Center (HMC) and Weill Cornell Medicine-Qatar.

## Results

Seroprevalence of SARS-CoV-2-IgG in this sample was estimated at 63.4% (449/708; 95% CI 59.8%-66.9%) using the BioMedomics assay and at 71.9% (509/708; 95% CI 68.5%-75.1%) using the Elecsys assay. Seroprevalence of SARS-CoV-2-IgM was estimated at 8.5% (60/708; 95% CI 6.6%-10.8%), measured only using the BioMedomics assay. Results of serological testing for each of the 708 participants are tabulated in Table S1 (Supplementary Information).

Among those seropositive in the Elecsys assay, optical density values (antibody titers) ranged between 1.0 and 150.0 with a median of 46.4 (Figure 1). There were 62 discordant results between the two assays. One specimen was seropositive in the BioMedomics assay, but seronegative in the Elecsys assay (0.1%). This specimen was drawn from a person who had no record of a prior SARS-CoV-2 diagnosis. However, there were 61 specimens seropositive in the Elecsys assay, but seronegative in the BioMedomics assay (8.6%).

**Figure 1.**
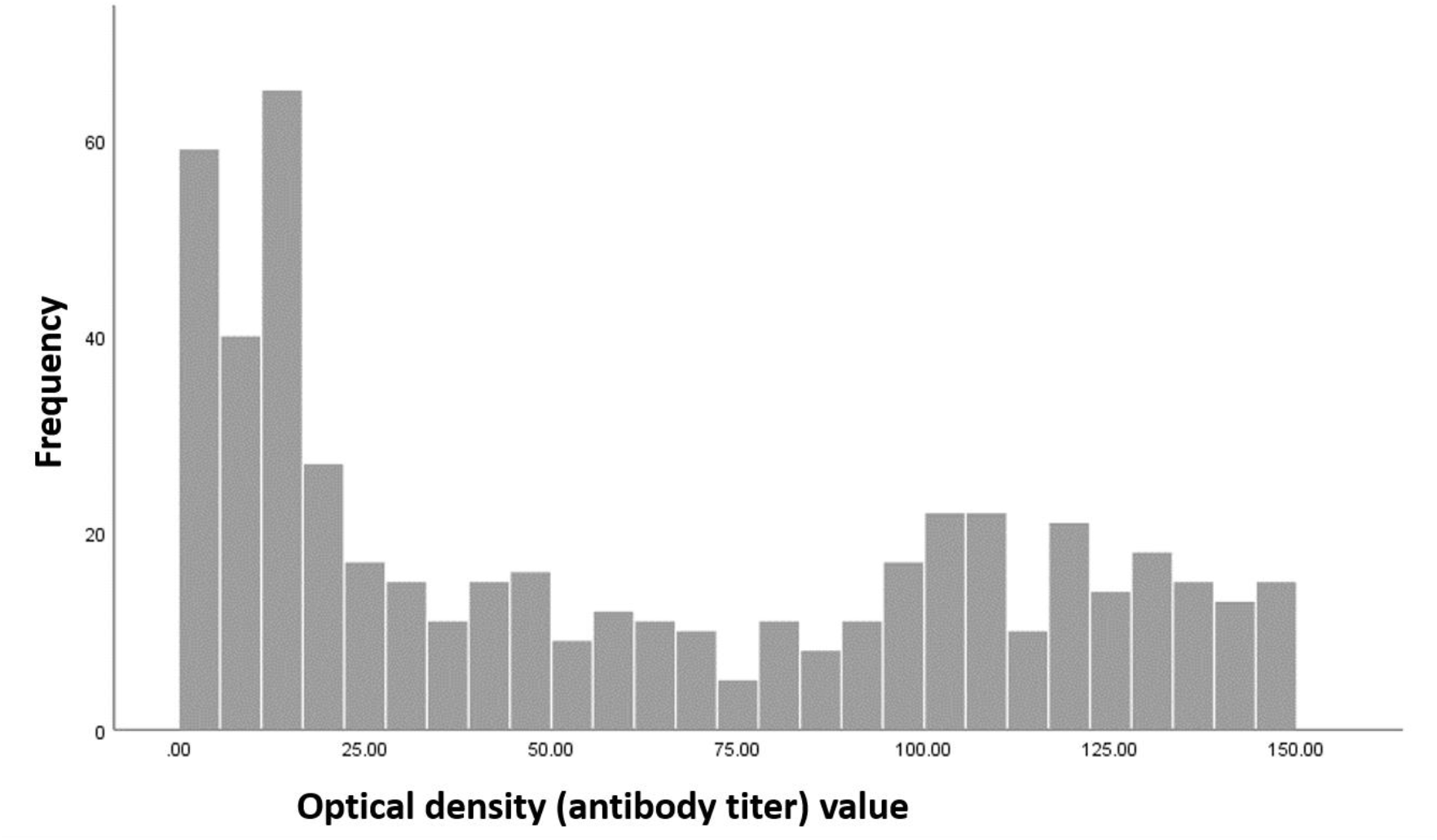
Distribution of optical density (antibody titer) values among 509 specimens that were seropositive in the Roche Elecsys Anti-SARS-CoV-2 test.

The percentage of individuals who had a PCR-confirmed SARS-CoV-2 diagnosis *prior* to the serological testing was 37.0% (262/708; 95% CI 33.5%-40.6%) (Table S1 of SI). PCR cycle threshold (Ct) values ranged between 12.5 and 38.3 with a median of 21.9. Among those seropositive in the BioMedomics assay, 53.2% (239/449; 95% CI 48.6%-57.8%) had received a prior PCR-positive result. Ct values ranged between 12.5 and 38.3 with a median of 21.8.

Among those seropositive in the Elecsys assay, 51.1% (260/509; 95% CI 46.8%-55.4%) had received a prior PCR-positive result. Ct values ranged between 12.5 and 38.3 with a median of 22.0. Among the 62 discordant specimens, 33.9% (21/62; 95% CI 23.3%-46.3%) had a prior PCR-positive result. Ct values ranged between 13.7 and 36.8 with a median of 22.9.

Among those with a prior PCR-positive result *and* were seropositive in the Elecsys assay, 91.9% (239/260; 95% CI 88.0%-94.7%) were seropositive in the BioMedomics assay. Two individuals were seronegative in both antibody assays, but had a prior PCR-confirmed SARS-CoV-2 diagnosis (Table S1 of SI). The first individual was diagnosed on May 21, 2020 (Ct value was 35.7), 151 days prior to the blood collection date. With the high Ct value, one cannot exclude the possibility of a PCR false positive result. The second individual was diagnosed on October 19, 2020 (Ct value was 19.7), only one day prior to the blood collection date, and thus the blood was probably drawn too early to detect development of antibodies.

Data on COVID-19 disease severity per the WHO classification [17] (derived from the national COVID-19 hospitalization database [19]) were available for 47 persons: 7 individuals were classified as mild, 19 were classified as moderate, 14 were classified as severe, and 7 were classified as critical (Table S1 of SI). For all other individuals no severity classification was conducted, due to absence of serious symptoms to require hospitalization and severity assessment, and thus the infection can be assumed to be asymptomatic or mild. No COVID-19 deaths were reported among study participants.

The positive, negative, and overall percent agreements between the two assays were estimated at 88.0% (95% CI 84.9%-90.6%), 99.5% (95% CI 97.2%-99.9%), and 91.2% (95% CI 88.9%-93.1%), respectively (Table 1). Cohen’s kappa statistic was estimated at 0.80 (95% CI 0.77-0.83) indicating excellent agreement between the two assays (Table 1).

**Table 1.** Concordance metrics between two SARS-CoV-2 antibody assays: the BioMedomics COVID-19 IgM/IgG Rapid Test and the Roche Elecsys Anti-SARS-CoV-2 including A) all negative and positive specimens, B) negative specimens and specimens with higher antibody titers (excluding specimens with Elecsys optical density values <10), and C) negative specimens and specimens with lower antibody titers (excluding specimens with Elecsys optical density values ≥10).

| A) |  | Roche Elecsys Anti-SARS-CoV-2 |  |  | Overall percent agreement | Positive percent agreement | Negative percent agreement | Cohen's kappa statistic |
| --- | --- | --- | --- | --- | --- | --- | --- | --- |
|  |  | Positive | Negative | Total | % (95% CI) | % (95% CI) | % (95% CI) | k (95% CI) |
| BioMedomics COVID-19 IgM/IgG Rapid Test | Positive | 448 | 1 | 449 | 91.2%<br>(88.9%-93.1%) | 88.0%<br>(84.9%-90.6%) | 99.5%<br>(97.2%-99.9%) | 0.80<br>(0.77-0.83) |
|  | Negative | 61 | 198 | 259 |  |  |  |  |
|  | Total | 509 | 199 | 708 |  |  |  |  |
| B) |  | Roche Elecsys Anti-SARS-CoV-2 |  |  | Overall percent agreement | Positive percent agreement | Negative percent agreement | Cohen's kappa statistic |
|  |  | Positive | Negative | Total | % (95% CI) | % (95% CI) | % (95% CI) | k (95% CI) |
| BioMedomics COVID-19 IgM/IgG Rapid Test | Positive | 385 | 1 | 386 | 93.9%<br>(91.7%-95.5%) | 91.2%<br>(88.2%-93.6%) | 99.5%<br>(97.2%-99.9%) | 0.87<br>(0.84-0.89) |
|  | Negative | 37 | 198 | 235 |  |  |  |  |
|  | Total | 422 | 199 | 621 |  |  |  |  |
| C) |  | Roche Elecsys Anti-SARS-CoV-2 |  |  | Overall percent agreement | Positive percent agreement | Negative percent agreement | Cohen's kappa statistic |
|  |  | Positive | Negative | Total | % (95% CI) | % (95% CI) | % (95% CI) | k (95% CI) |
| BioMedomics COVID-19 IgM/IgG Rapid Test | Positive | 63 | 1 | 64 | 91.3%<br>(87.4%-94.0%) | 72.4%<br>(62.2%-80.7%) | 99.5%<br>(97.2%-99.9%) | 0.78<br>(0.73-0.83) |
|  | Negative | 24 | 198 | 222 |  |  |  |  |
|  | Total | 87 | 199 | 286 |  |  |  |  |

Including only specimens with higher antibody titers in the comparison (i.e., excluding specimens with low Elecsys optical density values <10), the positive, negative, and overall percent agreements between the two assays were estimated at 91.2% (95% CI 88.2%-93.6%), 99.5% (95% CI 97.2%-99.9%), and 93.9% (95% CI 91.7%-95.5%), respectively (Table 1). Cohen’s kappa statistic was estimated at 0.87 (95% CI 0.84-0.89) indicating excellent and superior agreement between the two assays compared to the result for the full sample (Table 1). The cutoff at Elecsys optical density value of 10 (to indicate lower antibody titers) was informed by the distribution of the optical density values (Figure 1).

Including only specimens with low antibody titers in the comparison (i.e. excluding specimens with Elecsys optical density values ≥10), positive, negative, and overall percent agreements between the two assays were estimated at 72.4% (95% CI 62.2%-80.7%), 99.5% (95% CI 97.2%-99.9%), and 91.3% (95% CI 87.4%-94.0%), respectively (Table 1). Cohen’s kappa statistic was estimated at 0.78 (95% CI 0.72-0.83) indicating excellent agreement between the two assays, but still an inferior agreement compared to the result for the subsample of higher antibody titers, and for the full sample (Table 1).

Multivariable logistic regression identified significant associations between seropositivity using the BioMedomics assay and both the Elecsys optical density value and the PCR Ct value or its absence (that is no prior PCR-confirmed infection), but no association with the severity of the infection (Table 2). The adjusted odds ratio (aOR) of seropositivity was 5.06 (95% CI 2.72-9.39, p<0.001) for those with higher antibody titers (Elecsys optical density values ≥10) compared to those with lower antibody titers (Elecsys optical density values <10), reflecting much higher agreement between the two measures for specimens with higher antibody titers. The aOR of seropositivity was 0.00 (95% CI 0.00-0.03, p<0.001) for those with a negative result in the Elecsys assay, indicating a very low likelihood of being negative in the Elecsys assay, but positive in the BioMedomics assay.

**Table 2.**
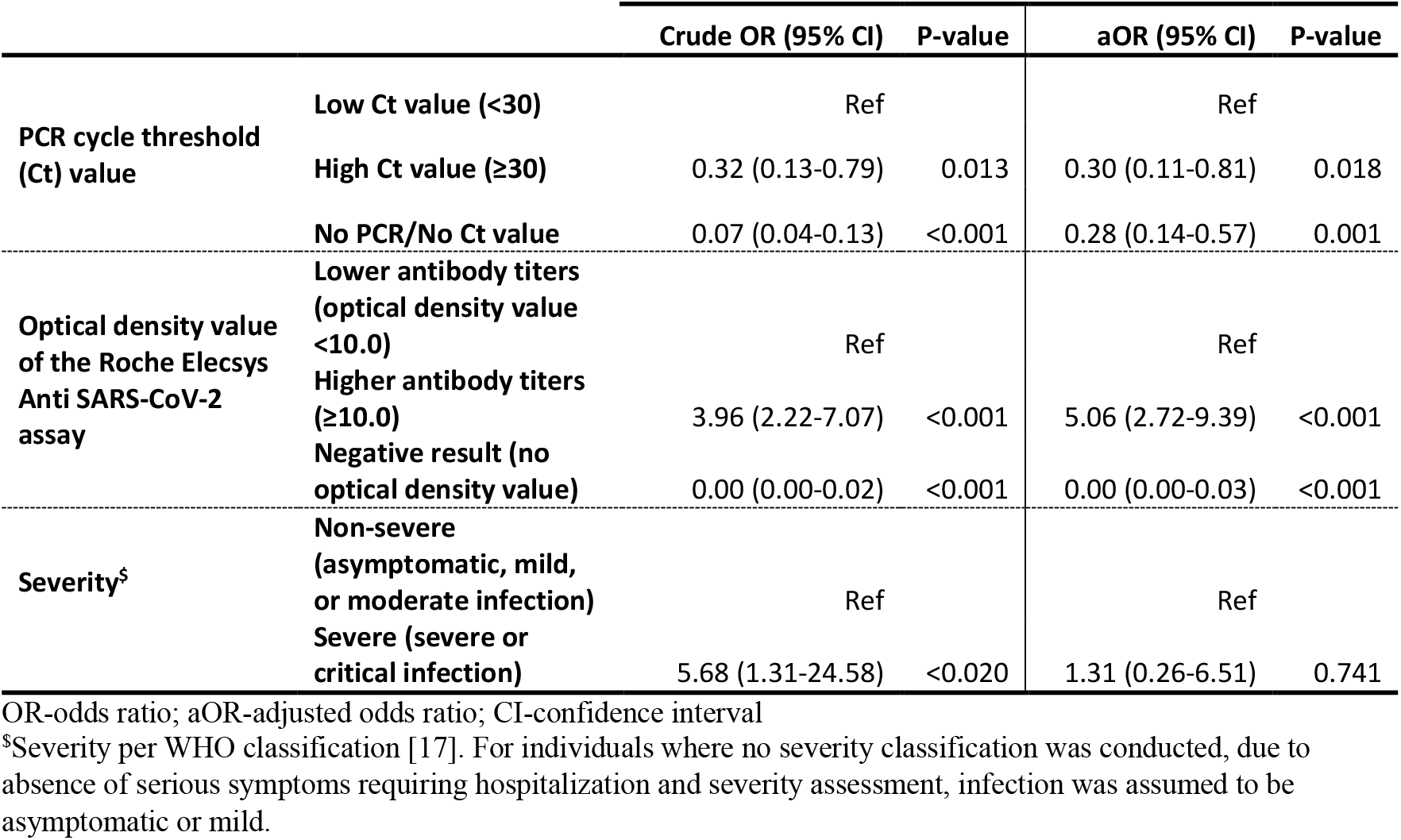
Results of univariable and multivariable logistic regression, assessing the association between seropositivity using the BioMedomics COVID-19 IgM/IgG Rapid Test and the following covariates: PCR Ct value, optical density from the Elecsys assay, and severity of infection.

The aOR of seropositivity was 0.30 (95% CI 0.11-0.81, p=0.018) for those with a Ct value ≥30 compared to those with a Ct value <30, possibly because of some PCR false positivity measures for those with higher Ct values. The aOR of seropositivity was 0.28 (95% CI 0.14-0.57, p=0.001) for those with no Ct value (that is no prior PCR-confirmed infection), reflecting a low likelihood of having been exposed to the infection if they were never diagnosed with the infection.

## Discussion

The above results document excellent performance of the BioMedomics rapid test in measuring detectable antibodies against SARS-CoV-2. An additional benefit is to help identify those with a PCR false positive result. The performance of this point-of-care test was also as expected for a rapid test: it was optimal when antibody titers were high, but less optimal when they were low. Only one test result could have been a false positive, suggesting very high specificity for this rapid test. The specimen was seronegative in the Elecsys assay and there was no record of a PCR-positive test for this individual.

These findings demonstrate the utility of this assay to assess past SARS-CoV-2 infection and potential seropositivity as a marker of immunity against infection, particularly considering the growing evidence indicating that natural infection elicits strong protection against reinfection that lasts for at least a few months [20–22]. These findings also support the concept of using rapid antibody testing for more efficient, cost-effective, and widescale serological testing to guide vaccine prioritization, or possible issuance of immunity passports.

This study has a limitation. The performance of the rapid assay was compared to a laboratory-based assay, the Roche Elecsys Anti SARS-CoV-2 platform (and to PCR testing), but not to a gold standard test of seropositivity, as such a test was not available. Having so said, the Elecsys assay is one of the most extensively used and investigated commercial platforms, having a specificity ≥99.8% [13–15] and a sensitivity ≥89% [3, 12, 15]. Therefore, it is not likely that this limitation could have affected the findings.

In conclusion, the BioMedomics rapid point-of-care test demonstrated excellent performance in measuring detectable antibodies against SARS-CoV-2, with better performance for specimens with higher antibody titers, demonstrating the utility of such assays in mass expansion of serological testing to guide public health responses and possible vaccine prioritization.

## Data Availability

All relevant data are available in the main text and supplementary material.

## Acknowledgments

We thank Her Excellency Dr. Hanan Al Kuwari, Minister of Public Health, for her vision, guidance, leadership, and support. We also thank Dr. Saad Al Kaabi, Chair of the System Wide Incident Command and Control (SWICC) Committee for the COVID-19 national healthcare response, for his leadership and analytical insights, and for his instrumental role in enacting data information systems that made these studies possible. We further express our appreciation to the SWICC Committee and members of the Scientific Reference and Research Taskforce (SRRT) for their informative input, scientific technical advice, and enriching discussions. We also thank Dr. Mariam Abdulmalik, CEO of the Primary Health Care Corporation and Chairperson of the Tactical Community Command Group on COVID-19, as well as members of this committee, for providing support to the teams that worked on field surveillance. We further thank Dr. Nahla Afifi, Director of Qatar Biobank (QBB), Ms. Tasneem Al-Hamad, Ms. Eiman Al-Khayat and the rest of the QBB team for their unwavering support in retrieving and analyzing samples and in compiling and generating databases for COVID-19 infection, as well as Dr. Asmaa Al-Thani, Chairperson of the Qatar Genome Programme Committee and Board Vice Chairperson of QBB, for her leadership of this effort. We also acknowledge the dedicated efforts of the Clinical Coding Team and the COVID-19 Mortality Review Team, both at Hamad Medical Corporation, and the Surveillance Team at the Ministry of Public Health. We thank Dr. Katharine J. Looker, medical writer from the University of Bristol, for reviewing the manuscript.

## Author contributions

PVC and LJA conceived and designed the study. PVC led the laboratory testing. SRD conducted the statistical analyses and co-wrote the first draft of the manuscript. HC contributed to the statistical analyses. LJA led the data analyses and co-wrote the first draft of the manuscript. All authors contributed to data collection and acquisition, database development, discussions and interpretation of the results, and to the writing of the manuscript. All authors have read and approved the final manuscript.

## Data and material availability

All relevant data are available in the main text and supplementary material.

## Supplementary Information

**Table S1.**
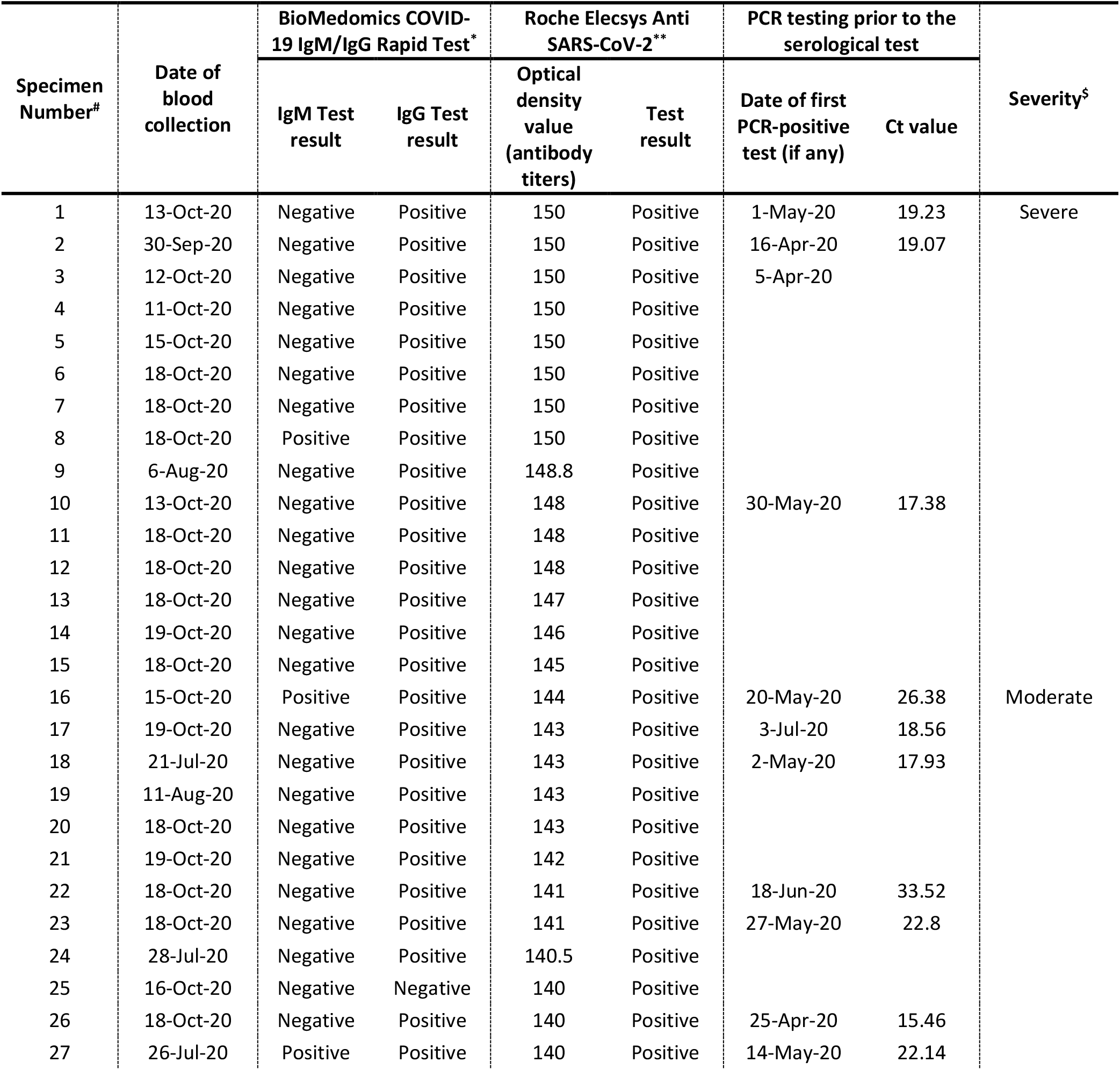
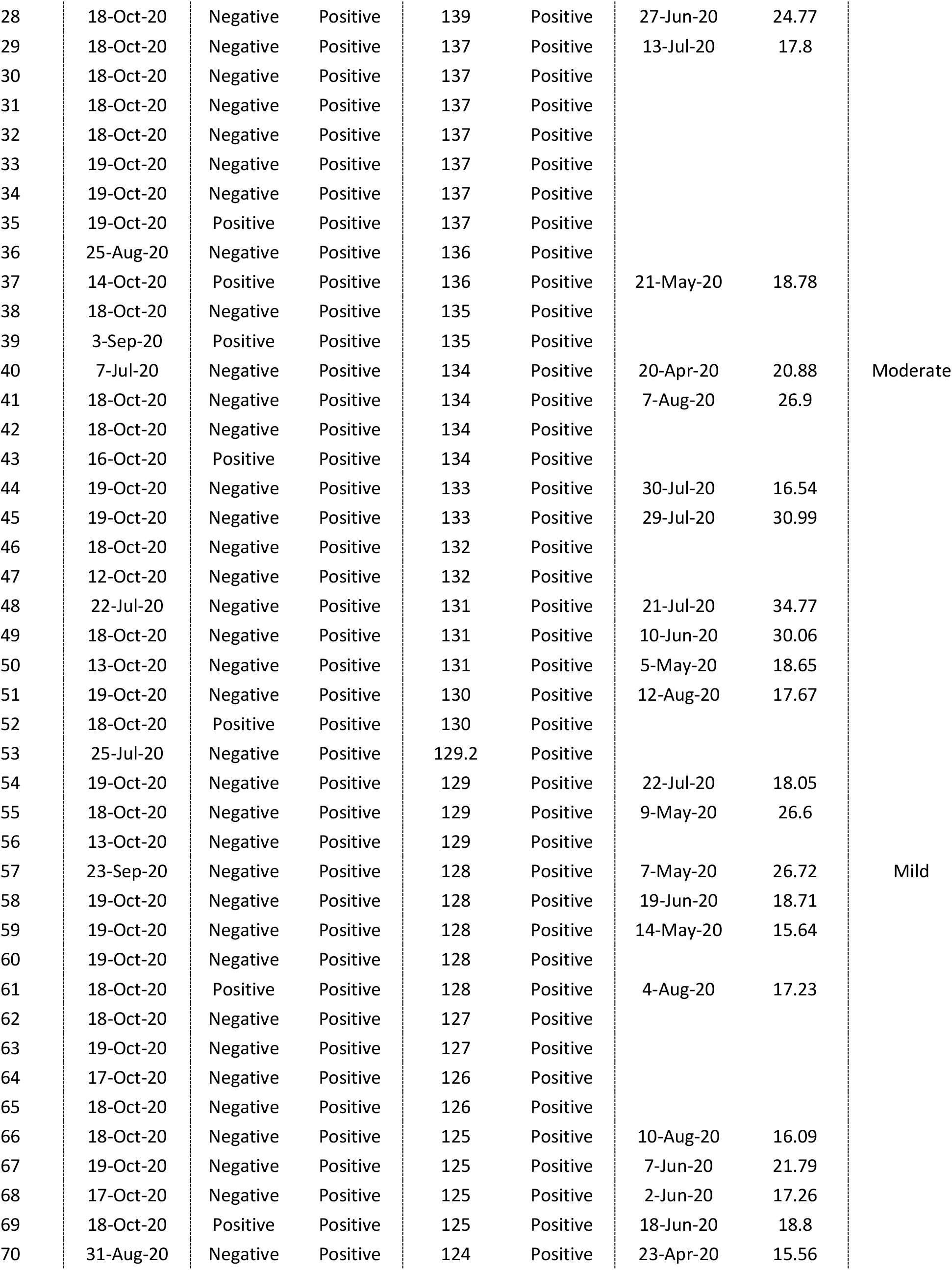
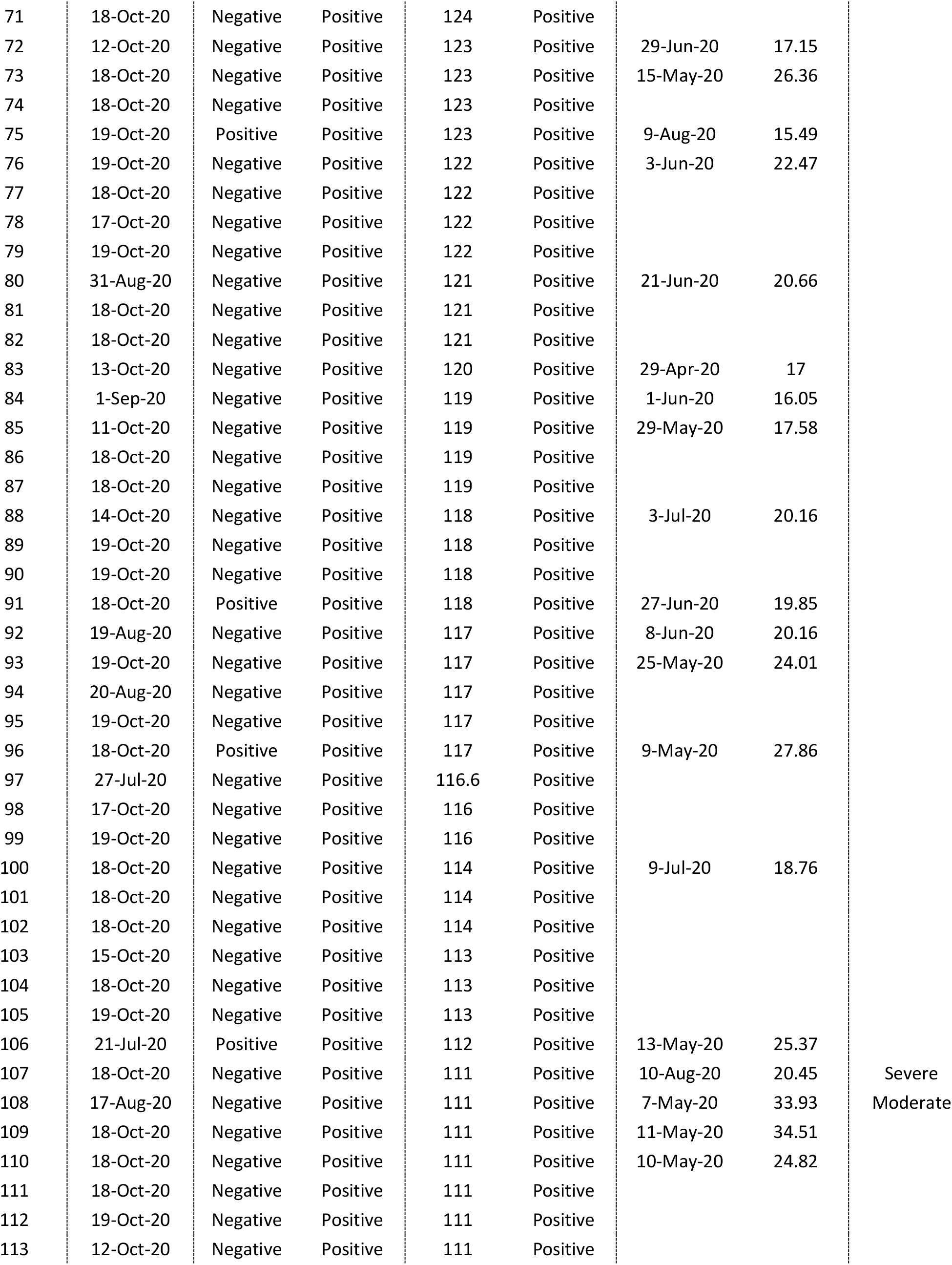
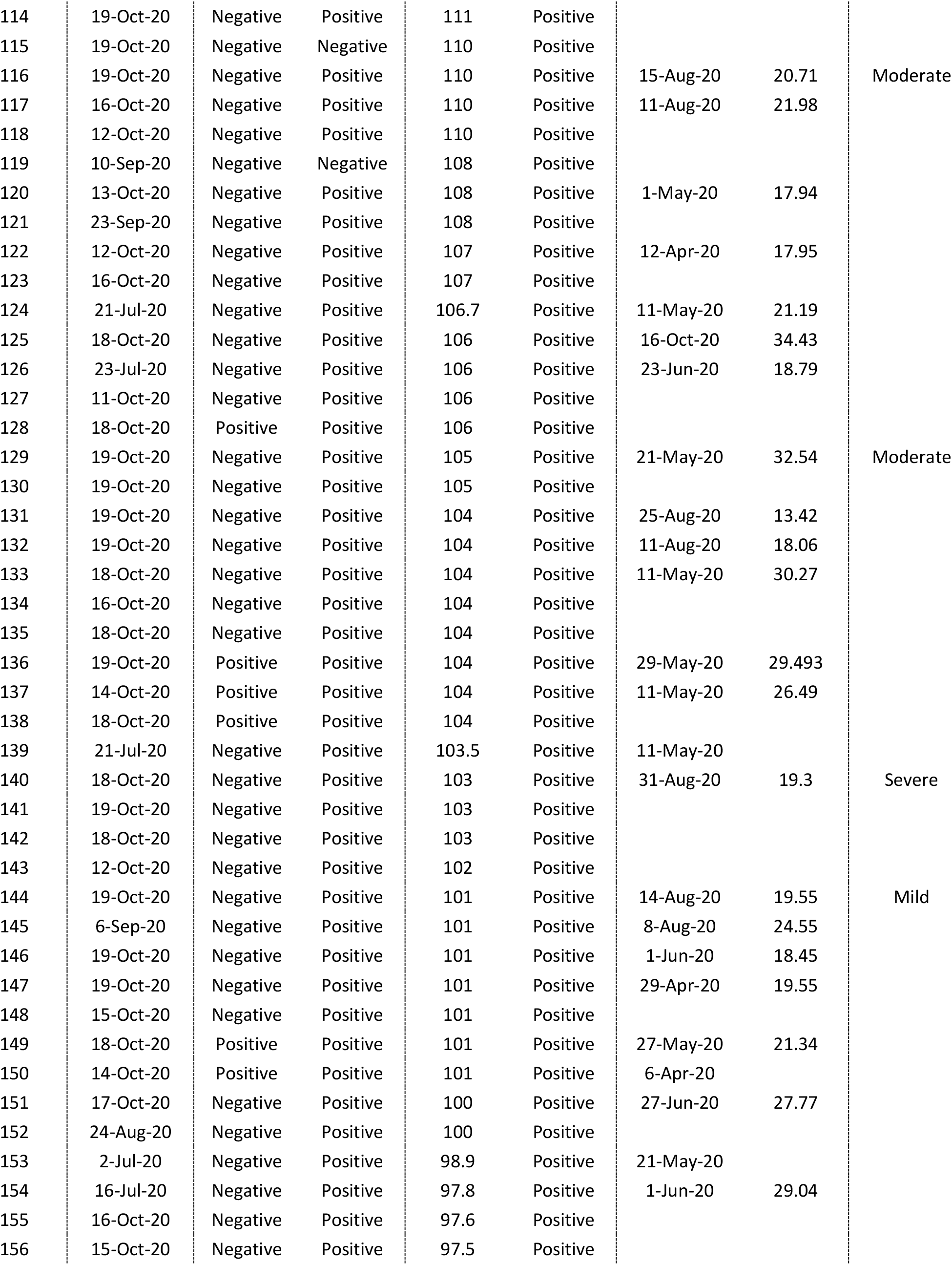
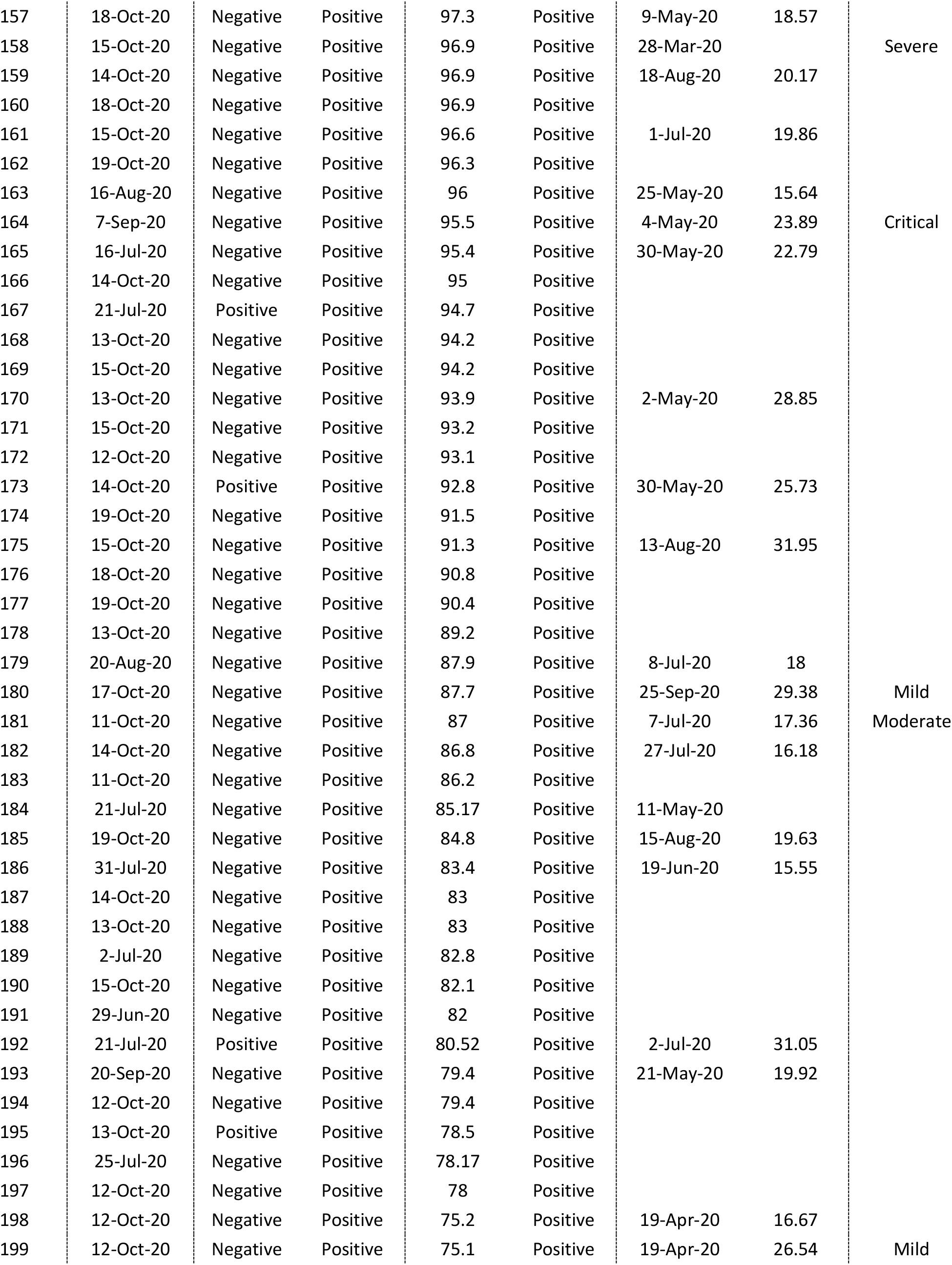
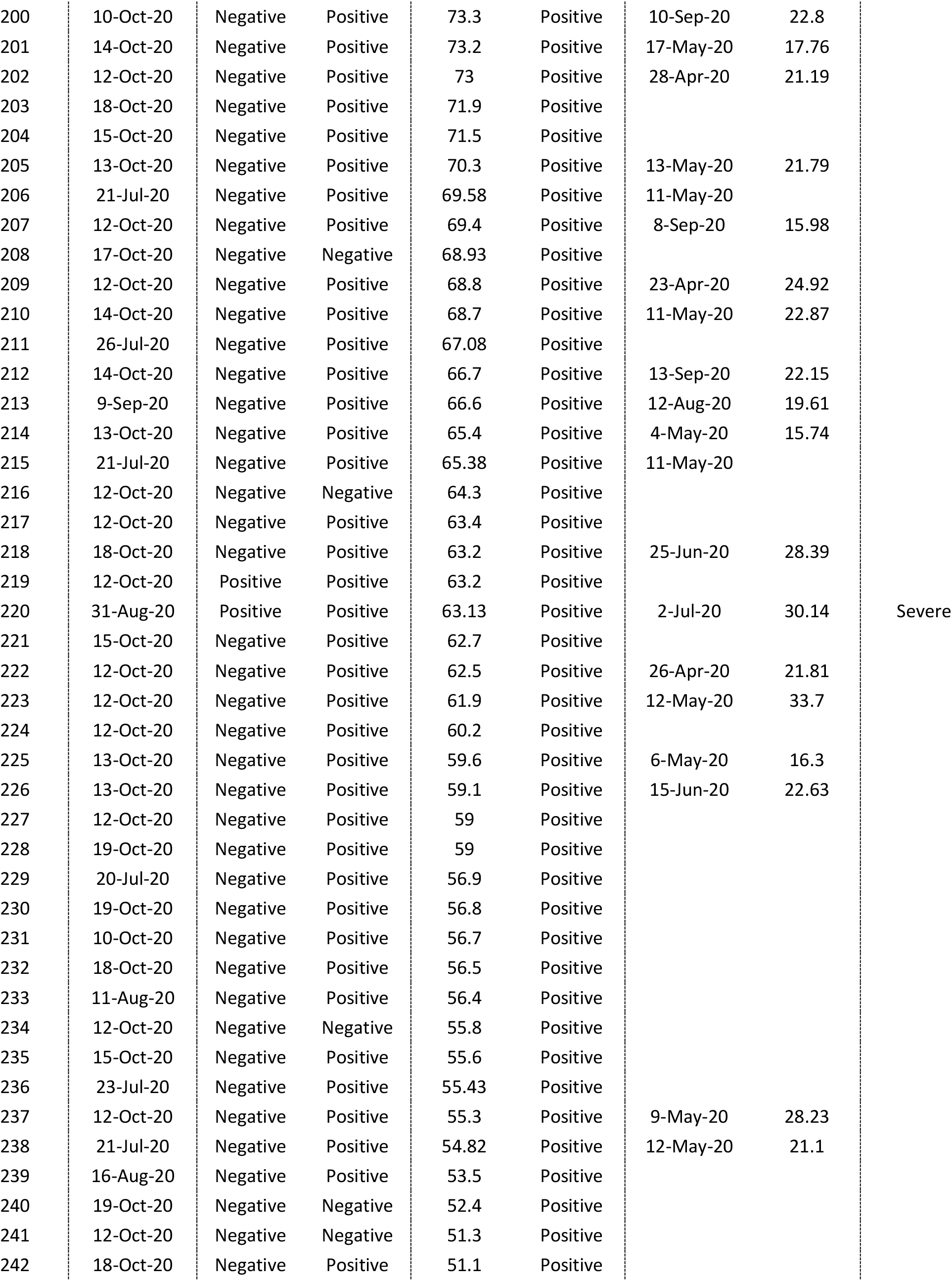
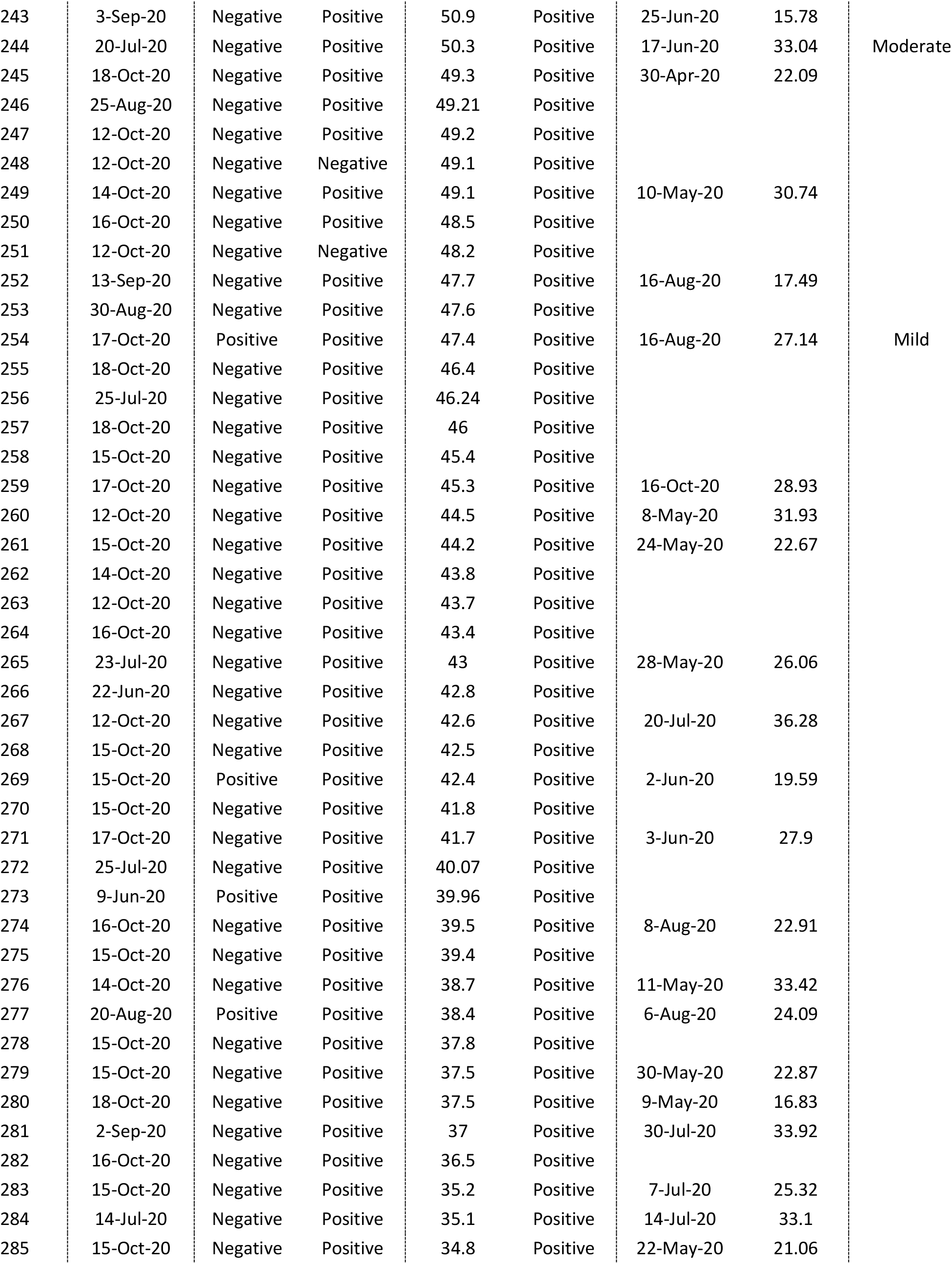
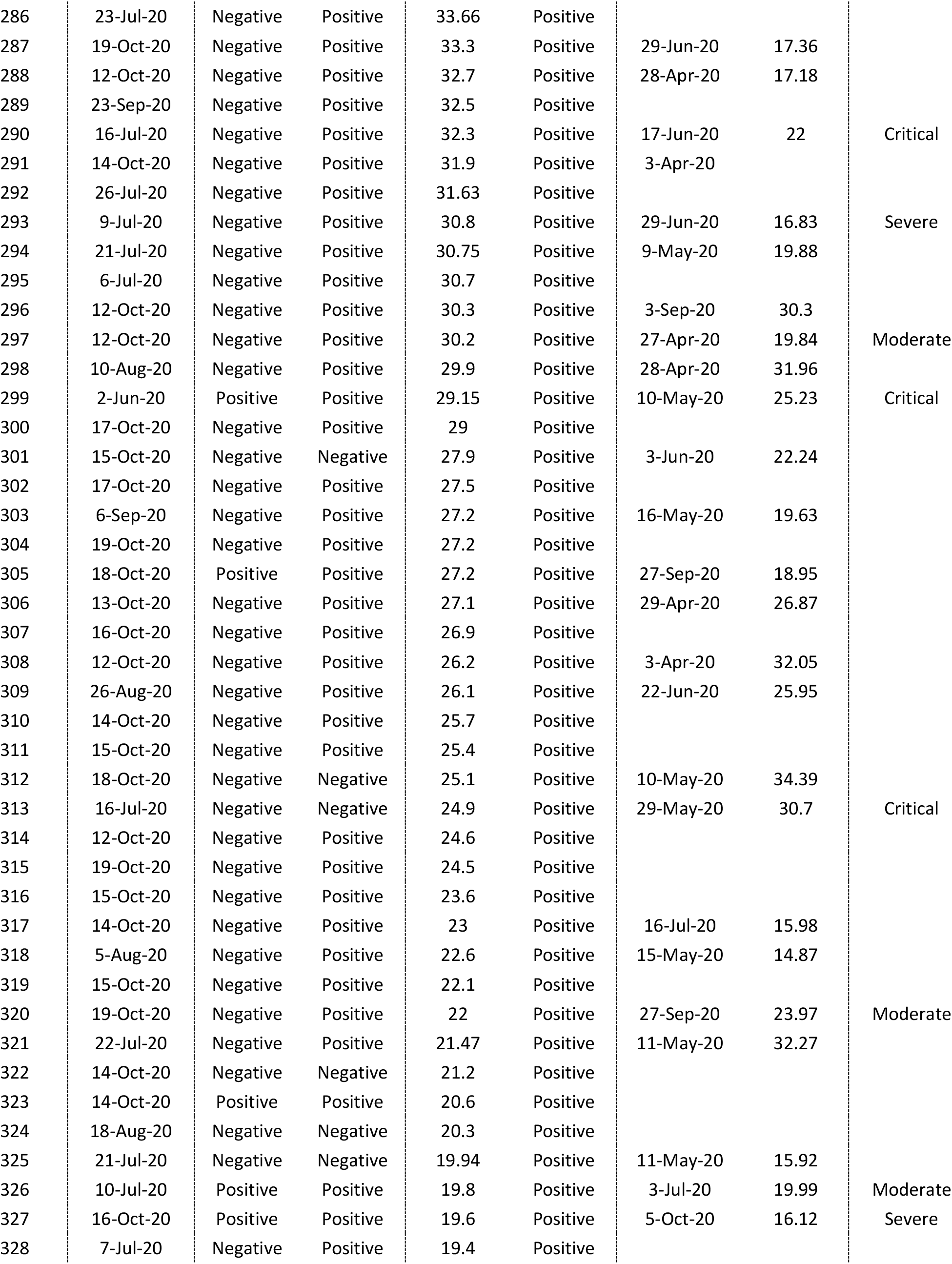
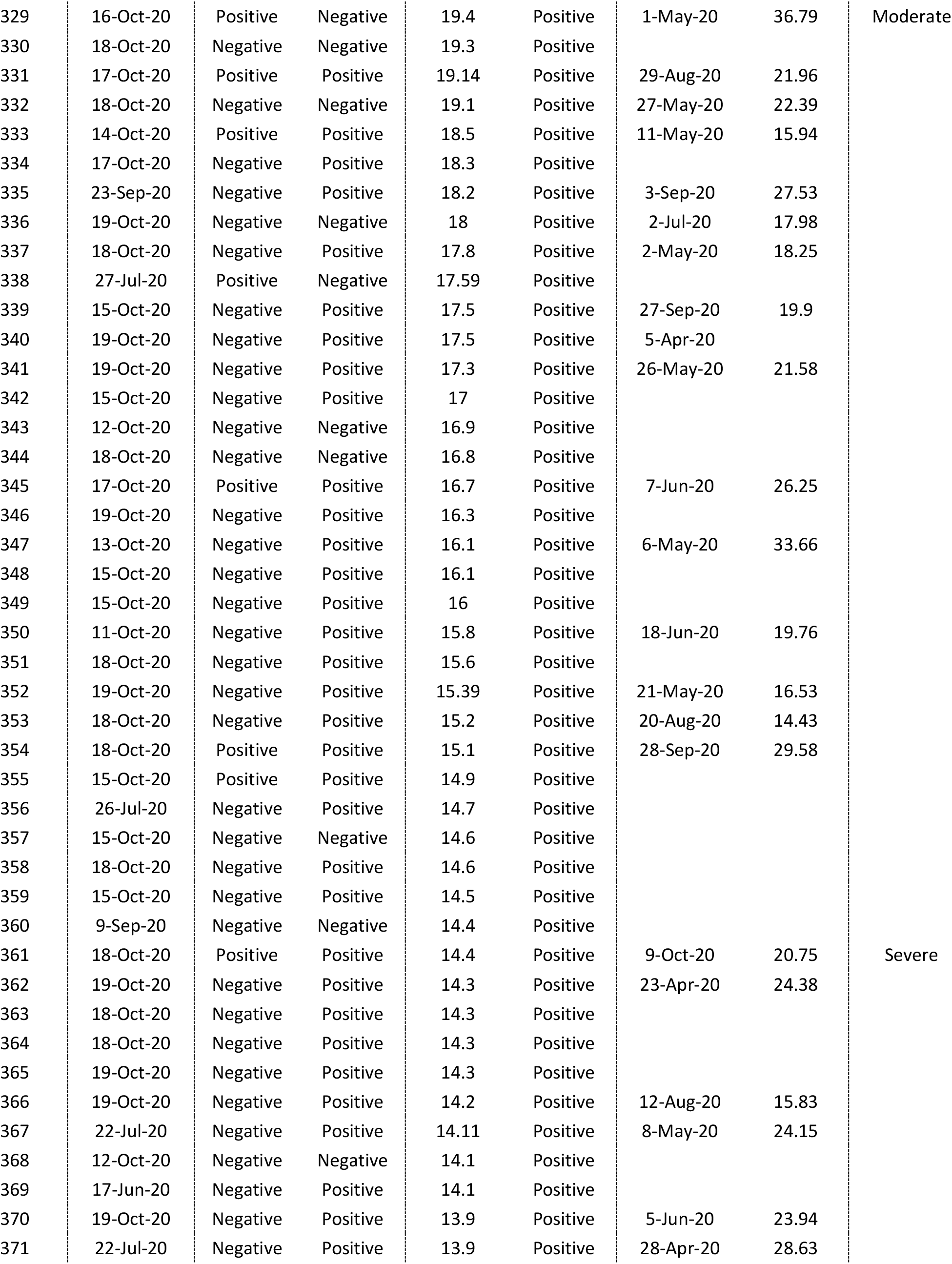
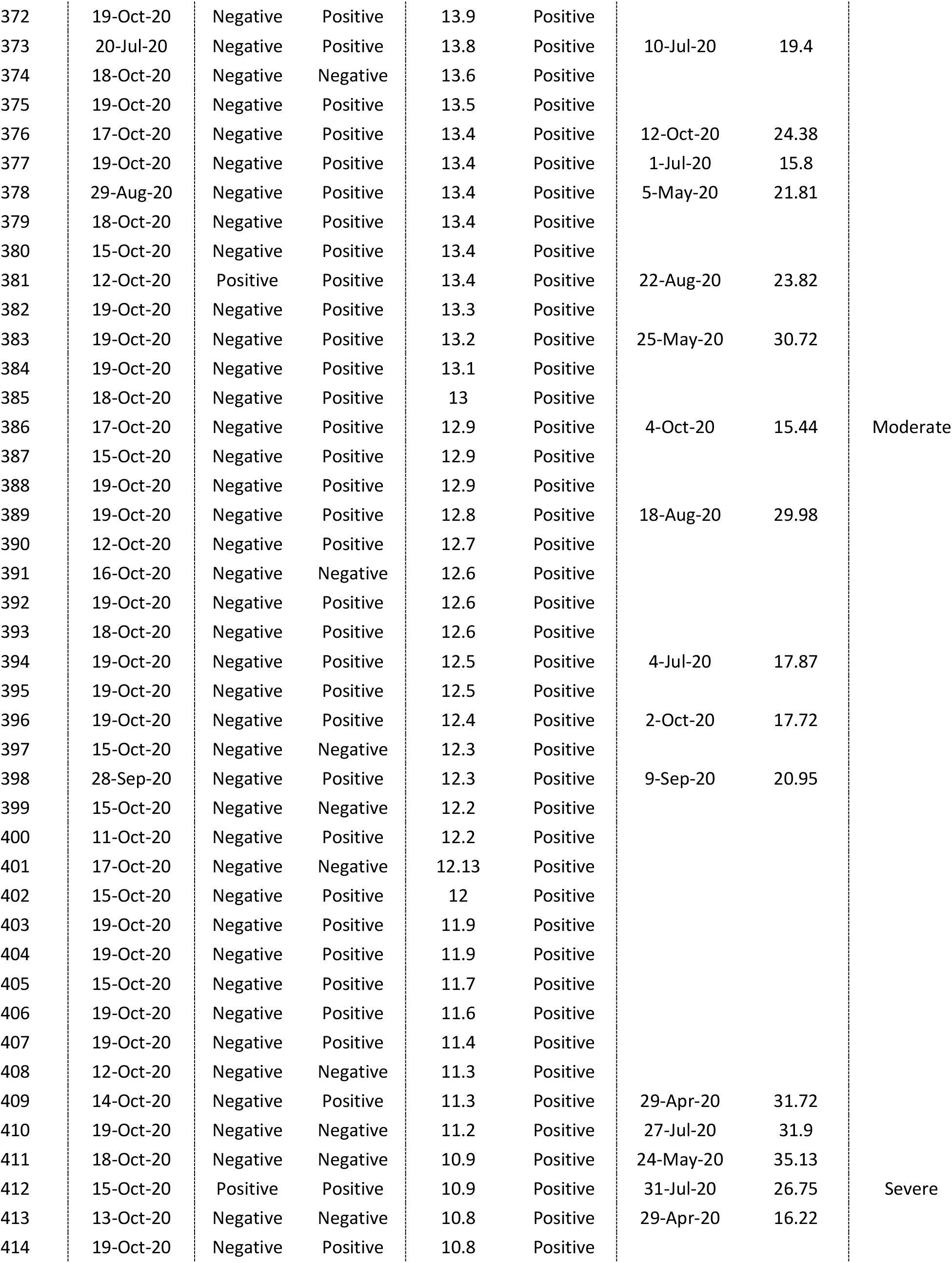
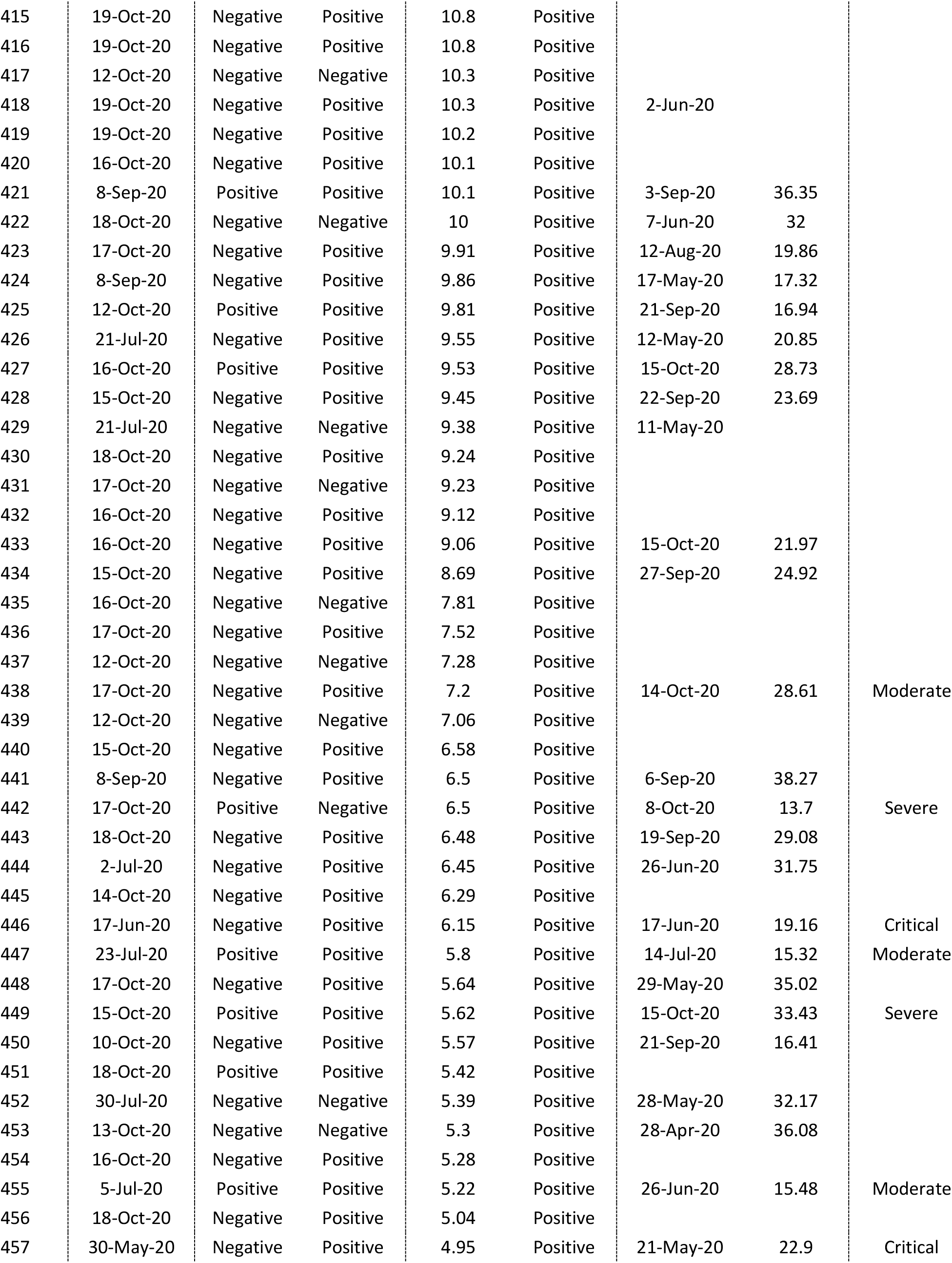
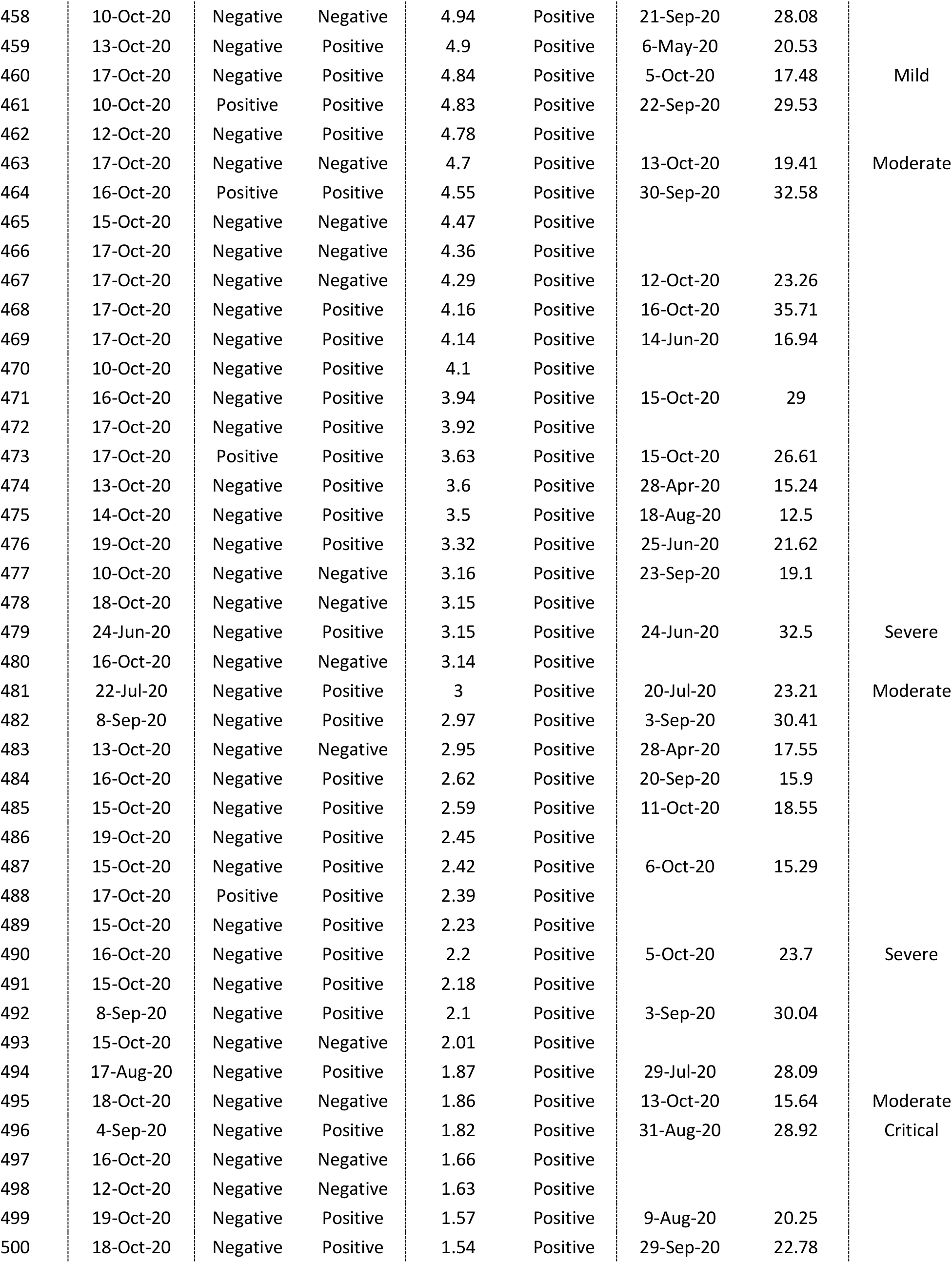
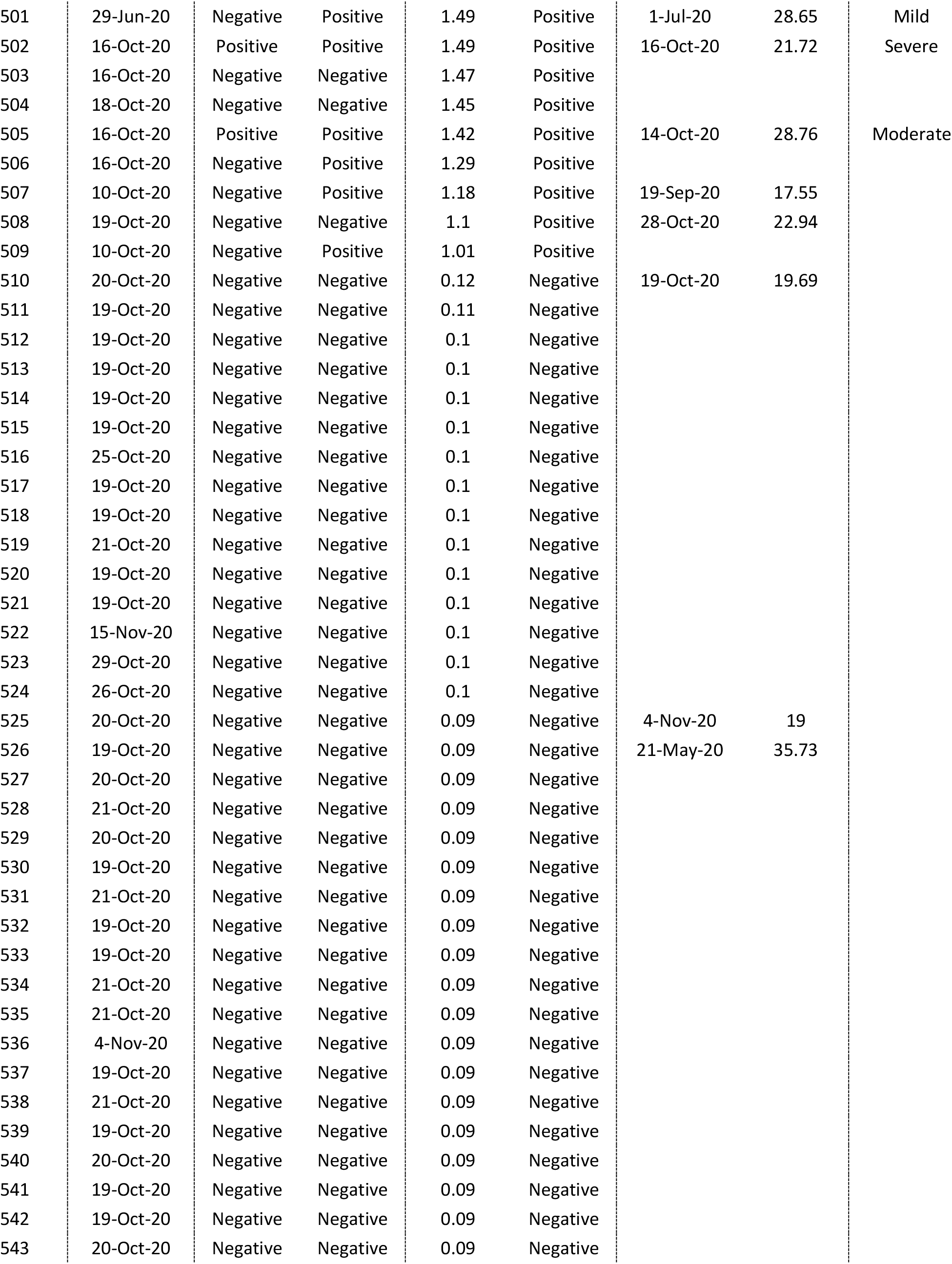
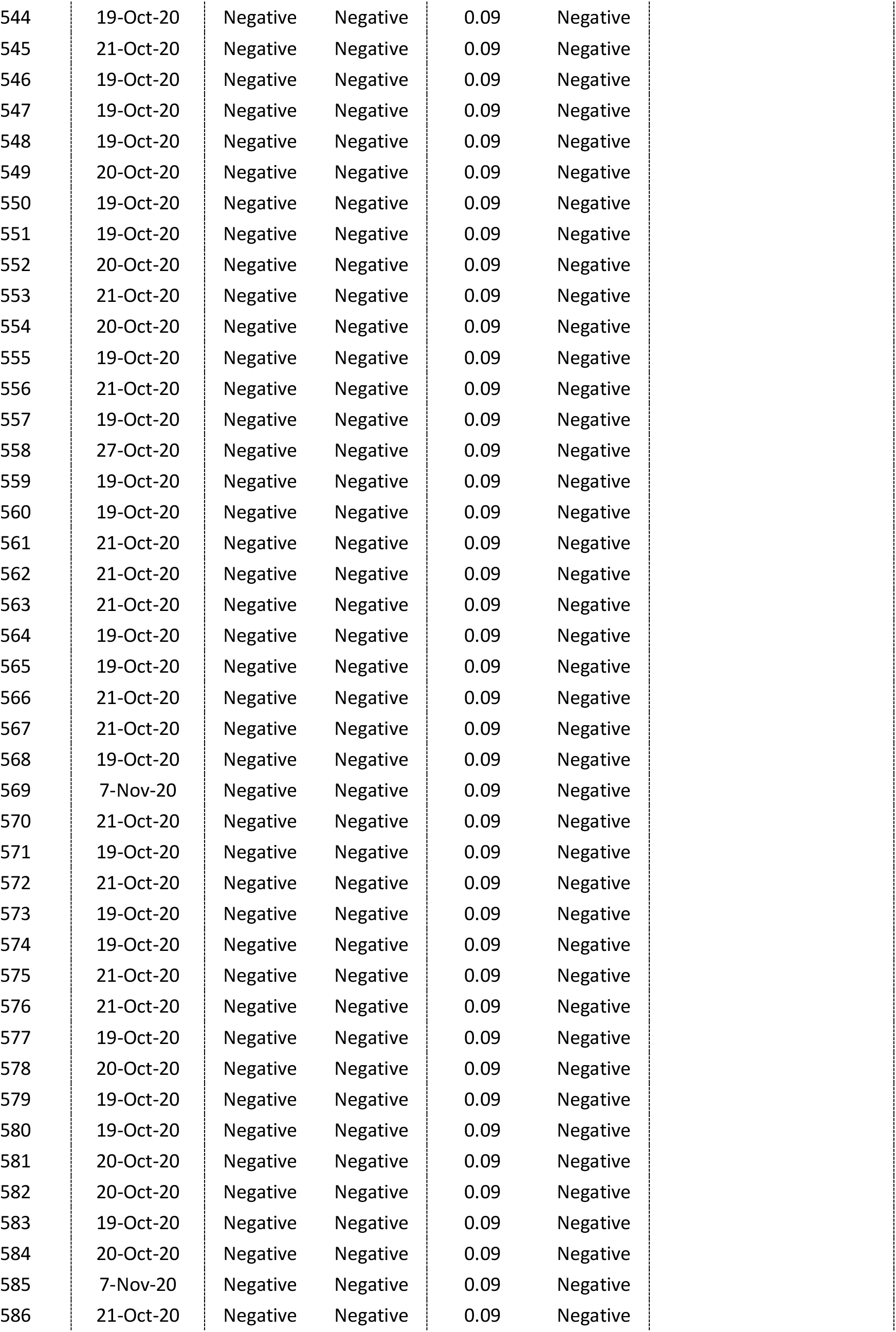
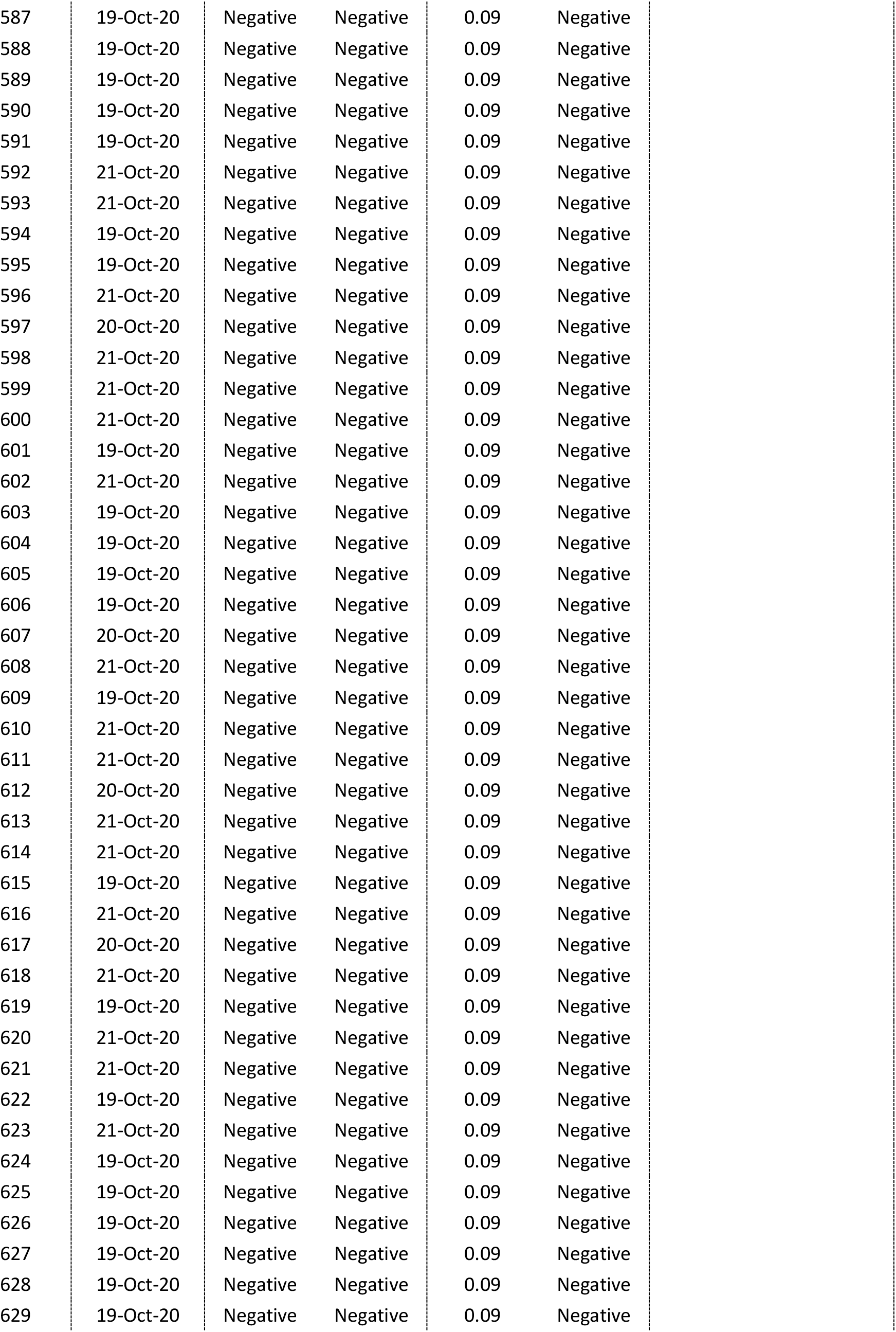
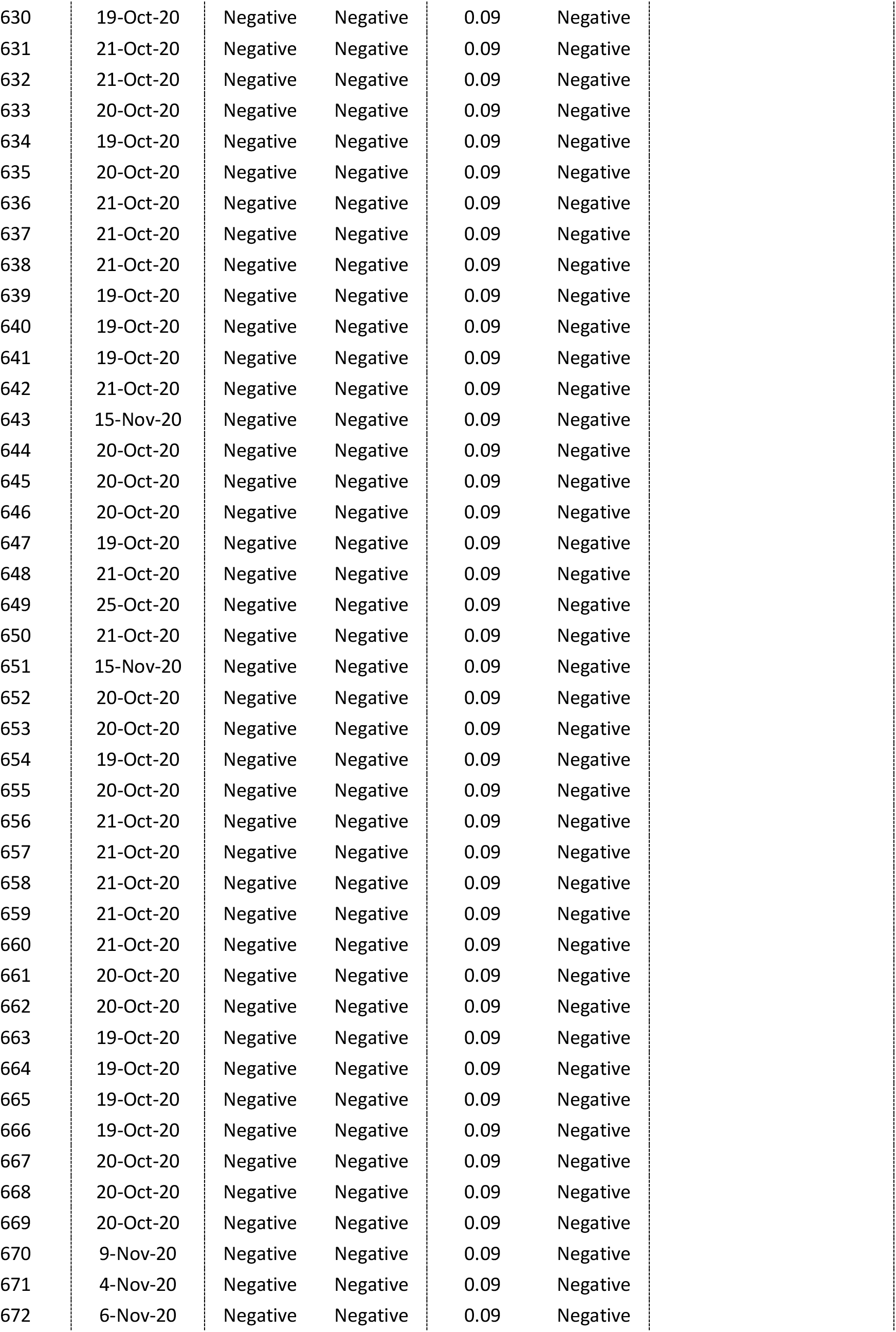
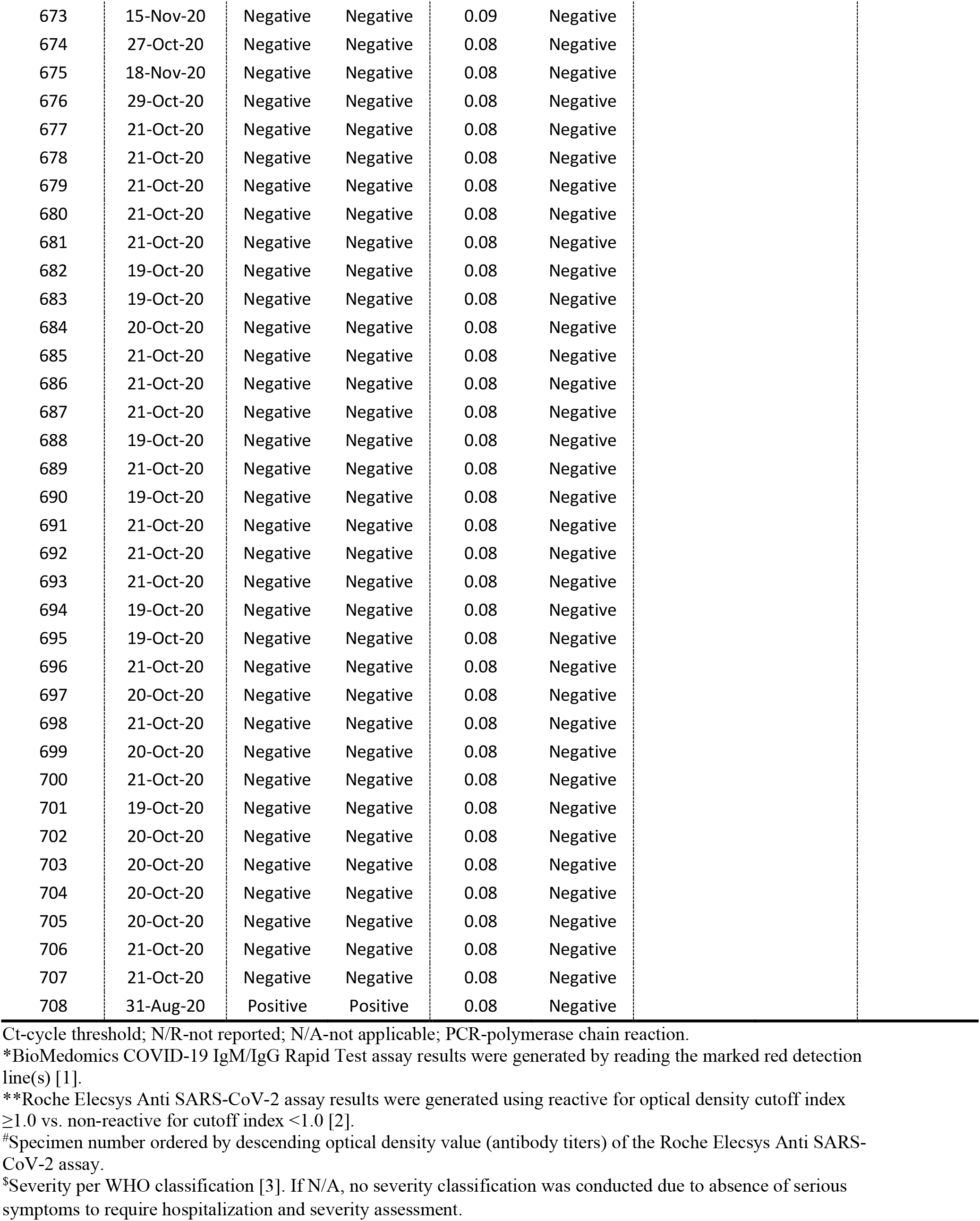
Characteristics of all specimens tested using the BioMedomics COVID-19 IgM/IgG Rapid Test and the Roche Elecsys Anti SARS-CoV-2 assay.

## References

1. De Walque D., Friedman J., Gatti R.V., Mattoo A. How two tests can help contain COVID-19 and revive the economy. Available from: http://documents.worldbank.org/curated/en/766471586360658318/pdf/How-Two-Tests-Can-Help-Contain-COVID-19-and-Revive-the-Economy.pdf. Accessed on April 16, 2020. Research & Policy Briefs, World Bank Malaysia Hub. 2020.

2. Nicola M, Alsafi Z, Sohrabi C, Kerwan A, Al-Jabir A, Iosifidis C, et al. The socio-economic implications of the coronavirus pandemic (COVID-19): A review. Int J Surg. 2020;78:185–93. doi: 10.1016/j.ijsu.2020.04.018. PubMed PMID: 32305533; PubMed Central PMCID: PMCPMC7162753.

3. Abu-Raddad LJ, Chemaitelly H, Ayoub HH, Al Kanaani Z, Al Khal A, Al Kuwari E, et al. Characterizing the Qatar advanced-phase SARS-CoV-2 epidemic. medRxiv. 2020:2020.07.16.20155317v2. doi: 10.1101/2020.07.16.20155317.

4. Ayoub HH, Chemaitelly H, Seedat S, Makhoul M, Al Kanaani Z, Al Khal A, et al. Mathematical modeling of the SARS-CoV-2 epidemic in Qatar and its impact on the national response to COVID-19. medRxiv. 2020:2020.11.08.20184663. doi: 10.1101/2020.11.08.20184663.

5. Al Kuwari HM, Abdul Rahim HF, Abu-Raddad LJ, Abou-Samra A-B, Al Kanaani Z, Al Khal A, et al. Epidemiological investigation of the first 5685 cases of SARS-CoV-2 infection in Qatar, 28 February–18 April 2020. BMJ Open. 2020;10(10):e040428. doi: 10.1136/bmjopen-2020-040428.

6. Al-Thani MH, Farag E, Bertollini R, Al Romaihi HE, Abdeen S, Abdelkarim A, et al. Seroprevalence of SARS-CoV-2 infection in the craft and manual worker population of Qatar. medRxiv. 2020:2020.11.24.20237719. doi: 10.1101/2020.11.24.20237719.

7. Coyle P., et al. National healthcare serological testing in the State of Qatar. under preparation.

8. Jeremijenko A, Chemaitelly H, Ayoub HH, Abdulla MAH, Abou-Samra AB, Al Ajmi JAAA, et al. Evidence for and level of herd immunity against SARS-CoV-2 infection: the ten-community study. medRxiv. 2020:2020.09.24.20200543. doi: 10.1101/2020.09.24.20200543.

9. Nasrallah GK, Dargham SR, Shurrab F, Al-Sadeq DW, Al-Jighefee H, Chemaitelly H, et al. Are commercial antibody assays substantially underestimating SARS-CoV-2 ever infection? An analysis on a population-based sample in a high exposure setting. medRxiv. 2020:2020.12.14.20248163. doi: 10.1101/2020.12.14.20248163.

10. Ayoub HH, Chemaitelly H, Makhoul M, Al Kanaani Z, Al Kuwari E, Butt AA, et al. Epidemiological impact of prioritizing SARS-CoV-2 vaccination by antibody status: Mathematical modeling analyses. MedRxiv. 2021:2021.01.10.21249382. doi: 10.1101/2021.01.10.21249382.

11. BioMedomics I. COVID-19 IgM/IgG Rapid Test. 2020. [November 1, 2020]. Available from: https://www.biomedomics.com/products/infectious-disease/covid-19-rt/.

12. Jahrsdörfer B, Kroschel J, Ludwig C, Corman VM, Schwarz T, Körper S, et al. Independent side-by-side validation and comparison of four serological platforms for SARS-CoV-2 antibody testing. The Journal of Infectious Diseases. 2020. doi: 10.1093/infdis/jiaa656.

13. The Roche Group. Roche’s COVID-19 antibody test receives FDA Emergency Use Authorization and is available in markets accepting the CE mark. 2020. [June 5, 2020]. Available from: https://www.roche.com/media/releases/med-cor-2020-05-03.htm.

14. Public Health England. Evaluation of Roche Elecsys AntiSARS-CoV-2 serology assay for the detection of anti-SARS-CoV-2 antibodies. Available from: https://assets.publishing.service.gov.uk/government/uploads/system/uploads/attachment_data/file/891598/Evaluation_of_Roche_Elecsys_anti_SARS_CoV_2_PHE_200610_v8.1_FINAL.pdf. Accessed on June 5, 2020. 2020.

15. Oved K, Olmer L, Shemer-Avni Y, Wolf T, Supino-Rosin L, Prajgrod G, et al. Multi-center nationwide comparison of seven serology assays reveals a SARS-CoV-2 non-responding seronegative subpopulation. EClinicalMedicine. 2020;29:100651. doi: 10.1016/j.eclinm.2020.100651. PubMed PMID: 33235985; PubMed Central PMCID: PMCPMC7676374.

16. Hamad Medical Corporation. National SARS-CoV-2 PCR testing, infection severity, and hospitalization database. 2020.

17. World Health Organization. Clinical management of COVID-19. Available from: https://www.who.int/publications-detail/clinical-management-of-covid-19. Accessed on: May 31st 2020. 2020.

18. Fleiss JL, Levin B, Paik MC. The Measurement of Interrater Agreement. Statistical Methods for Rates and Proportions: Wiley; 2013. p. 598–626.

19. Hamad Medical Corporation. National SARS-CoV-2 PCR and antibody testing, infection severity, and hospitalization database. 2020.

20. Abu-Raddad LJ, Chemaitelly H, Malek JA, Ahmed AA, Mohamoud YA, Younuskunju S, et al. Assessment of the risk of SARS-CoV-2 reinfection in an intense re-exposure setting. medRxiv. 2020:2020.08.24.20179457. doi: 10.1101/2020.08.24.20179457.

21. Lumley SF, O’Donnell D, Stoesser NE, Matthews PC, Howarth A, Hatch SB, et al. Antibody Status and Incidence of SARS-CoV-2 Infection in Health Care Workers. N Engl J Med. 2020. Epub 2020/12/29. doi: 10.1056/NEJMoa2034545. PubMed PMID: 33369366.

22. Abu-Raddad LJ, Chemaitelly H, Coyle P, Malek JA, Ahmed AA, Mohamoud YA, et al. SARS-CoV-2 reinfection in a cohort of 43,000 antibody-positive individuals followed for up to 35 weeks. MedRxiv. 2021:2021.01.15.21249731. doi: 10.1101/2021.01.15.21249731.

## References

1. BioMedomics I. COVID-19 IgM/IgG Rapid Test. 2020. [November 1, 2020]. Available from: https://www.biomedomics.com/products/infectious-disease/covid-19-rt/.

2. The Roche Group. Roche’s COVID-19 antibody test receives FDA Emergency Use Authorization and is available in markets accepting the CE mark. 2020. [June 5, 2020]. Available from: https://www.roche.com/media/releases/med-cor-2020-05-03.htm.

3. World Health Organization. Clinical management of COVID-19. Available from: https://www.who.int/publications-detail/clinical-management-of-covid-19. Accessed on: May 31st 2020. 2020.

